# Cause-Specific Excess Mortality in the US During the COVID-19 Pandemic

**DOI:** 10.1101/2024.03.05.24303783

**Authors:** Ekaterina Degtiareva, Andrea M. Tilstra, Jonas Schöley, Ridhi Kashyap, Jennifer B. Dowd

**Affiliations:** Department of Sociology, University of Oxford; St Catherine’s College, University of Oxford; Leverhulme Centre for Demographic Science, Nuffield Department of Population Health, University of Oxford; Nuffield College, University of Oxford; Max Planck Institute for Demographic Research

**Keywords:** COVID-19, excess mortality, United States

## Abstract

The COVID-19 pandemic was a significant shock to United States mortality, and it is important to understand how the pandemic impacted other causes of death. We estimated monthly excess mortality in the US by cause of death, age, and sex, for official deaths at ages 15 and older. Data come from the CDC Wonder Multiple Cause of Death database. We used a compositionally robust Generalized Additive Model (GAM) to estimate expected mortality counts in March 2020-December 2022 for eight causes of death: accidents, cardiovascular diseases, cancer, diabetes, influenza and pneumonia, substance-related (drugs and alcohol), suicide, and residual (including COVID-19 related deaths). Analyses were stratified by sex and 15-year age groups from 15-29 to 75+. Excess mortality was calculated as observed deaths minus expected deaths. From March 2020 to December 2022, we estimated 1 298 763 total excess deaths (95% CI: 1 226 542 to 1 370 804). While there were fewer deaths than expected due to some causes like flu/pneumonia and suicide, the largest number of excess deaths, excluding COVID-19, were attributed to cardiovascular diseases (115 765 deaths, 95% CI: 98 697 to 133 783) and substance use (86 637 deaths, 95% CI: 79 273 to 93 690). Percent excess substance-related mortality was high across all ages, while percent excess from cardiovascular diseases was highest at midlife ages. Some of these excess cardiovascular deaths were likely due to undercounted COVID-19 deaths, but others may reflect indirect impacts of the pandemic on healthcare utilization or longer-term effects of COVID-19 infections.

**SIGNIFICANCE STATEMENT:** The COVID-19 pandemic increased mortality directly due to COVID-19 deaths, but also changed the pattern of deaths due to other causes in the United States. We estimated excess cause-specific mortality in the US and present several findings. We estimated nearly 1.3 million total excess deaths in the US from March 2020 to December 2022. Deaths from suicide and influenza and pneumonia were lower than expected based on previous trends, but deaths due to cardiovascular diseases, diabetes, accidents, and substance-related causes (drug and alcohol) were higher. Cancer deaths were generally unchanged. By quantifying both the direct and indirect effects of the COVID-19 pandemic on US mortality, we highlight areas of on-going vulnerability in the US.

## INTRODUCTION

The COVID-19 pandemic was a significant shock to the mortality landscape in the United States, with a confirmed COVID-19 death toll of 1 172 229 as of January 20th, 2024^1^. Early estimates of US excess mortality were higher than official COVID-19 deaths^2,3^, suggesting undercounted COVID-19 mortality and/or increases in other causes of death during the pandemic. COVID-19 may also have longer-term impacts on mortality risk after acute infection or due to disruptions in health care delivery, resulting in persistent excess mortality^4^.

Understanding the ripple effects of the pandemic on US mortality is critical. Historically, the impact of pandemics was felt even after the acute phase of widespread infection, via the long-term health impacts of infection and the indirect social and economic consequences of the pandemic and policy responses^5^. In the context of the COVID-19 pandemic, there is evidence of interruptions in healthcare systems^6^, social isolation^7^, economic difficulties^8^, and increases in psychological stress^9^. It was feared that these social shocks might increase mortality from non-COVID causes, such as deaths by suicide because of the psychological strain or cancer deaths from missed screenings and treatments.

The US entered the pandemic with more vulnerable population health compared to peer nations, including an ongoing opioid crisis^10^, increasing midlife mortality^11^, and overall declining or stalling life expectancy^12^. The pandemic exacerbated these trends, as the US experienced larger losses of life expectancy than many of its peers^13–15^. Declines were greatest among males^13–15^ and Black and Hispanic Americans^16,17^.

Some existing work has explored changes in US pandemic mortality for specific causes of death^18–20^. This evidence suggests that the pandemic contributed to increases in deaths from cardiovascular diseases^21^ and Alzheimer’s disease^22^, reduced suicide rates^23^ contrasted by an uptick in substance abuse deaths^24^, and a notable displacement effect of other seasonal respiratory diseases^25^. These studies looked at individual causes in isolation and for different time periods, but a comprehensive study of changes in cause-specific mortality in the US during the pandemic has not yet been done. We use a compositional regression approach to estimate cause-specific excess mortality in the US from March 2020 through December 2022. Our method ensures coherency between total and cause-specific estimated excess deaths and explicitly addresses the dependency in deaths across causes. We provide comprehensive estimates of both the direct and indirect impact of the COVID-19 pandemic on US mortality to better understand how these trends might persist and how medicine, public health, and public policy might best restore and improve population health post-pandemic.

## RESULTS

We estimated 1 298 763 total excess deaths (95% CI: 1 226 542 to 1 370 804) between March 2020 and December 2022 in the United States (Table 1). The pattern of excess deaths fluctuated throughout the period, as depicted in Figure 1 (and zoomed in eFigure 1). Over this time official COVID-19 deaths (solid purple line) tracked alongside excess deaths in the residual death category (shaded peach area). From May 2022 onward, official COVID-19 deaths accounted for a smaller proportion of residual excess deaths, with the emergence of persistent non-COVID-19 excess in the absence of big COVID-19 surges.

**Figure 1.**
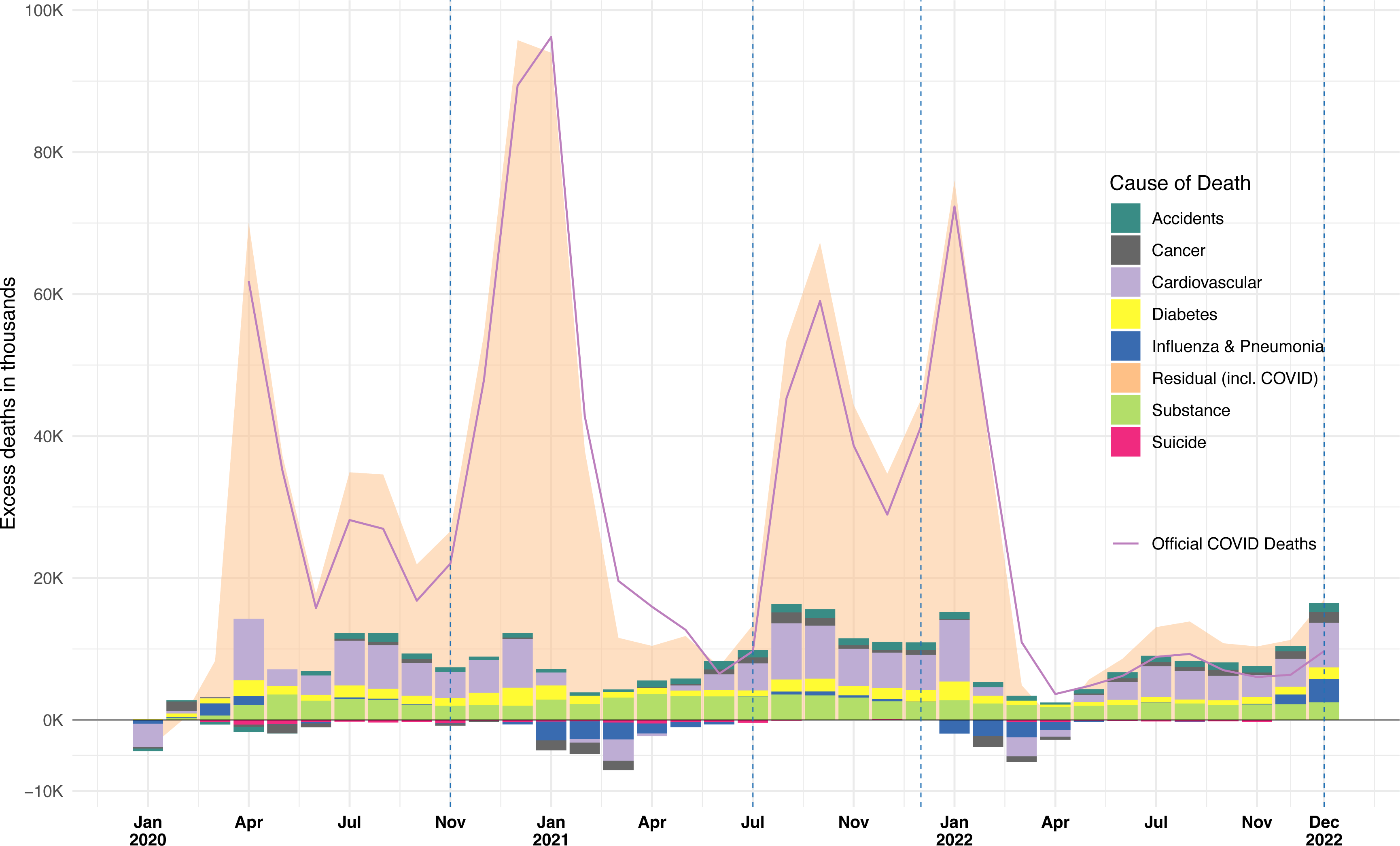
Absolute excess mortality by cause of death, United States, Jan 2020 to Dec 2022. Notes: Data come from CDC Wonder and authors’ own calculations. Dashed lines indicate periods of dominant SARS-CoV-2 variants.

**Table 1.** Total cumulative excess mortality by cause of death and age group, United States females (Panel A) and males (Panel B), March 2020 to December 2022.

| Panel A. Females |  |  |  |  |  |  |  |  |  |  |  |
| --- | --- | --- | --- | --- | --- | --- | --- | --- | --- | --- | --- |
|  | 15-29 |  | 30-44 |  | 45-59 |  | 60-74 |  | 75+ |  | Total |
|  | Median | (95% CI) | Median | (95% CI) | Median | (95% CI) | Median | (95% CI) | Median | (95% CI) | Median (95% CI) |
| Residual | 4 568 | (4 360; 4 815) | 16 998 | (16 603; 17 334) | 51 210 | (50 353; 51 996) | 132 11 | (129 659; 134 423) | 267 121 | (257 465; 278 089) | 472 015 (458 440; 486 657) |
| Accidents | 1 538 | (1 348; 1 728) | 1 622 | (1 382; 1 873) | 903 | (698; 1 094) | 436 | (127; 732) | 2 831 | (2 235; 3 456) | 7 330 (5 790; 8 883) |
| Cancer | -75 | (-195; 39) | 663 | (331; 1 012) | -729 | (-1 453; -20) | 1 204 | (15; 2 444) | 2 921 | (1 233; 4 719) | 3 984 (-69; 8 194) |
| CVD | 148 | (51; 241) | 2 683 | (2 448; 2 903) | 6 865 | (6 247; 7 460) | 16 762 | (15 144; 18 385) | 29 884 | (24 235; 36 653) | 56 342 (48 125; 65 642) |
| Diabetes | 236 | (181; 287) | 709 | (601; 816) | 2 926 | (2 712; 3 152) | 5 390 | (5 027; 5 745) | 7 745 | (7 282; 8 206) | 17 006 (15 803; 18 206) |
| Influenza | -169 | (-298; -69) | -414 | (-598; -251) | -832 | (-1 178; -510) | -949 | (-1 733; -217) | -4 392 | (-6 514; -2 486) | -6 756 (-10 321; -3 533) |
| Substance | 2 106 | (1 620; 2 526) | 9 000 | (8 307; 9 637) | 9 310 | (8 768; 9 791) | 5 132 | (4 743; 5 531) | 1 212 | (996; 1 408) | 26 760 (24 434; 28 893) |
| Suicide | -125 | (-293; 32) | -138 | (-329; 57) | -1 037 | (-1 203; -875) | -543 | (-697; -401) | 55 | (-11; 117) | -1 788 (-2 533; -1 070) |
| Total | 8 227 | (6 774; 9 599) | 31 123 | (28 745; 33 381) | 68 616 | (64 944; 72 088) | 159 550 | (152 285; 166 642) | 307 377 | (286 921; 330 162) | 574 893 (539 669; 611 872) |
| Panel B. Males |  |  |  |  |  |  |  |  |  |  |  |
|  | 15-29 |  | 30-44 |  | 45-59 |  | 60-74 |  | 75+ |  | Total |
|  | Median | (95% CI) | Median | (95% CI) | Median | (95% CI) | Median | (95% CI) | Median | (95% CI) | Median (95% CI) |
| Residual | 10 892 | (10 350; 11 401) | 32 589 | (32 122; 33 042) | 87 485 | (86 755; 88 204) | 190 267 | (188 191; 192 535) | 249 197 | (242 521; 254 915) | 570 430 (559 939; 580 097) |
| Accidents | 4 953 | (4 497; 5 368) | 5 982 | (5 595; 6 327) | 2 348 | (1 919; 2 810) | 2 606 | (2 091; 3 097) | 1 809 | (1 189; 2 347) | 17 698 (15 291; 19 949) |
| Cancer | 130 | (9; 258) | 1 338 | (1 103; 1 575) | 956 | (375; 1 540) | -405 | (-2 053; 1 174) | -2 466 | (-3 893; -1 202) | -447 (-4 459; 3 345) |
| CVD | 491 | (353; 630) | 5 672 | (5 297; 6 039) | 14 038 | (13 107; 14 963) | 21 811 | (19 568; 24 163) | 17 411 | (12 247; 22 346) | 59 423 (50 572; 68 141) |
| Diabetes | 381 | (317; 441) | 1 224 | (1 076; 1 364) | 5 560 | (5 275; 5 830) | 8 867 | (8 403; 9 309) | 8 288 | (7 825; 8 746) | 24 320 (22 896; 25 690) |
| Influenza | -75 | (-159; -8) | -26 | (-154; 104) | -696 | (-1 108; -339) | 79 | (-756; 788) | -1 082 | (-2 738; 416) | -1 800 (-4 915; 961) |
| Substance | 7 118 | (6 065; 8 127) | 20 041 | (18 145; 21 823) | 21 408 | (20 265; 22 583) | 10 587 | (9 912; 11 255) | 723 | (452; 1 009) | 59 877 (54 839; 64 797) |
| Suicide | -1 477 | (-1 919; -1 078) | 111 | (-222; 462) | -2 333 | (-2 681; -1 996) | -2 008 | (-2 343; -1 712) | 76 | (-125; 276) | -5 631 (-7 290; -4 048) |
| Total | 22 413 | (19 513; 25 139) | 66 931 | (62 962; 70 736) | 128 766 | (123 907; 133 595) | 231 804 | (223 013; 240 609) | 273 956 | (257 478; 288 853) | 723 870 (686 873; 758 932) |

Notes: Data come from CDC Wonder and authors' own calculations.

Besides official COVID-19 deaths, several causes of death showed excess above what was expected over this period. Cardiovascular diseases stood out as the leading cause of non-COVID-19-attributed excess deaths, with a peak of 8 682 excess deaths in January 2022 during the Omicron wave. Across the whole period, there were 115 765 excess deaths attributed to CVD (95% CI: 98 697 to 133 783 [eTable 2]). Excess deaths associated with substance use, accidents, and diabetes were consistently elevated over the period, totaling 86 637 (95% CI: 79 273 to 93 690), 25 028 (95% CI: 21 081 to 28 832) and 41 326 (95% CI: 38 699 to 43 896) excess deaths over the period [eTable 2], respectively. Fewer than expected deaths (negative excess) were seen for some causes including deaths due to influenza and pneumonia (-8 556 [95% CI: -15 236 to -2 572]) and suicide (-8 556 [95% CI: -15 236 to -2 572]) (eTable 2). Overall, there were no big differences in observed versus expected cancer deaths, but there were some significant elevations for older women coupled with fewer than expected cancer deaths in older men (Table 1).

Figure 2 shows a different view of excess mortality for each cause, with predicted deaths (represented by lines) and observed deaths (represented by dots). These panels highlight the seasonal patterns for different causes of death and the accurate fit of our model to pre-pandemic deaths. For example, there are consistently fewer influenza and pneumonia-induced deaths during the summer months, and fewer suicides during the colder months. During the time of the pandemic, these patterns were even more exaggerated in the US, with suicide and influenza/pneumonia deaths below expected levels. In contrast, substance-related deaths exhibited less seasonality, even at baseline (prior to March 2020), and had consistently high excess across the pandemic starting in April 2020. Using relative P-scores, eFigure 2 shows that CVD deaths fluctuated between 5% and 10% excess each month of the pandemic. Relative increases for substance-related deaths were substantially higher, around 30% higher than expected each month.

**Figure 2.**
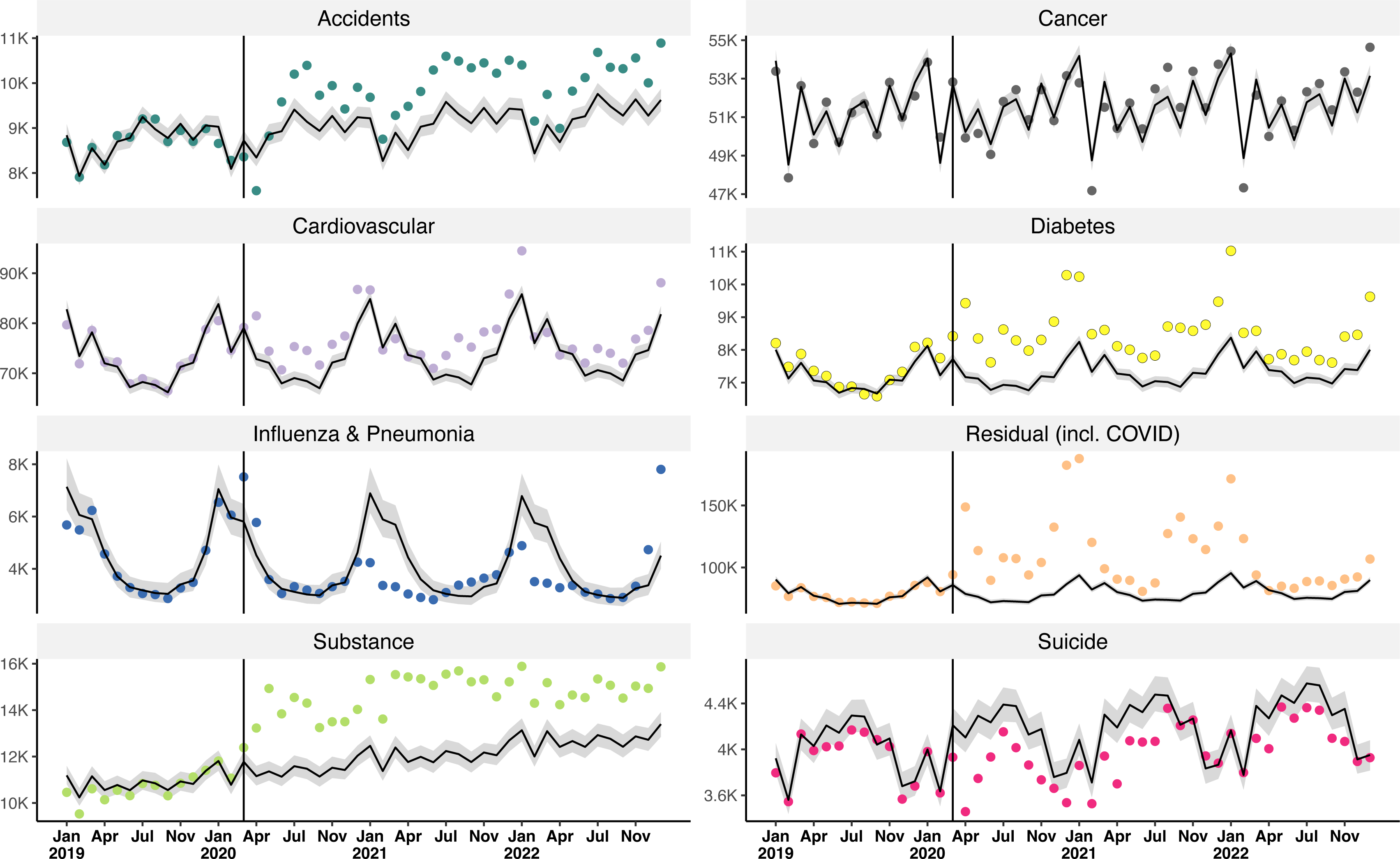
Observed and forecasted deaths by cause, US, January 2019 to December 2022. Notes: Data come from CDC Wonder and authors’ own calculations. Dots represent observed values. Solid line represents forecasted values, with shaded 95% prediction intervals.

Cumulative P-scores by cause and age-sex group are shown in Figure 3, where orange indicates a higher cumulative P-score and positive excess mortality, and blue indicates more negative cumulative P-score and negative excess mortality (fewer deaths than expected). All confidence intervals are reported in eTable 3. Despite the higher absolute numbers of excess deaths from CVD at older ages (Table 1), 30-44 year-olds experienced the greatest relative increase in CVD deaths (females: 17.52% [95% CI: 15.75 to 19.24]; males: 18.55% [95% CI: 17.12 to 19.99], Figure 3 and eTable 3). Relative increases were especially high for substance-related deaths, with deaths 25% higher than expected for males aged 15-59. There were fewer deaths than expected for influenza and pneumonia deaths across all age-sex groups except for males 60-74 for whom there was little change. Consistent with results in Figures 1 and 2, cancer deaths showed little relative cumulative excess, except for males aged 30-44 (8.5% [95% CI: 6.91 to 10.16], Figure 3 and eTable 3). Deaths due to accidents dropped for the first month of the pandemic, but soon climbed to above-expected levels with high relative excess, especially at younger ages (15-44) (19.63% [95% CI: 17.5 to 21.63] for males and 17.02% [95% CI: 14.61 to 19.53] for females age 15-29, Figure 3 and eTable 3) compared to older ages (60-74) (2.03% [95% CI: 0.58 to 3.46] for males and 5.89% [95% CI: 4.67 to 7.08] for females, Figure 3 and eTable 3).

**Figure 3.**
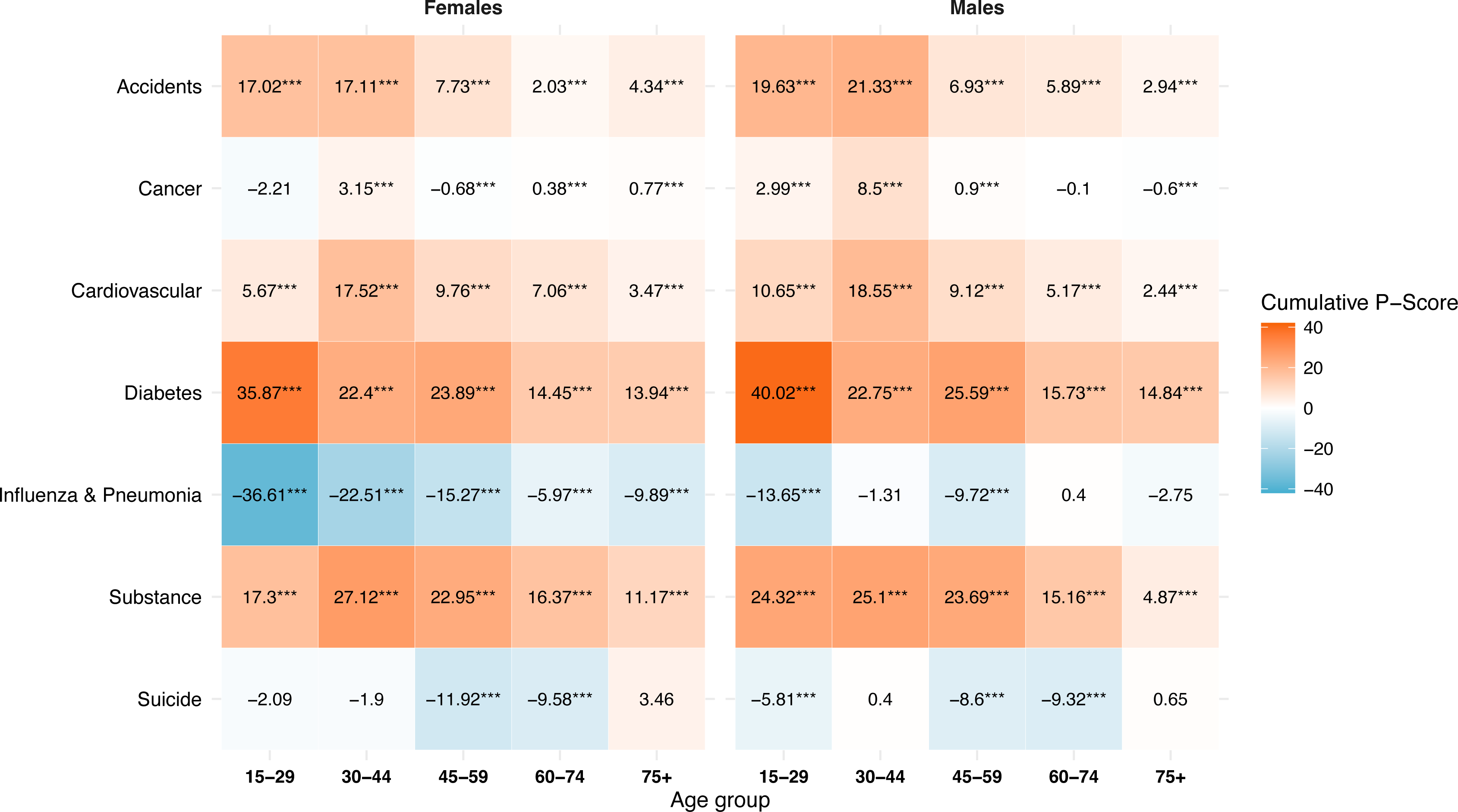
Cumulative relative mortality (percent excess), by cause and strata, US, March 2020 to December 2022. Notes: Data come from CDC Wonder and authors’ own calculations.

## DISCUSSION

The mortality burden of the COVID-19 pandemic in the US remains staggering, despite how normalized these numbers have become. The US accumulated 1.3 million excess deaths by the end of 2022, a number that would have seemed unfathomable in early 2020. These deaths represent almost 0.4% of the population, exceeding the US death rate due to World War II^26^. US life expectancy losses in 2020 and 2021 were among the worst in high-income countries and have not yet recovered to pre-pandemic levels^14,27^. Besides the direct effects of the SARS-CoV-2 virus, the significant social and economic disruptions of the pandemic also affected population health indirectly. While previous studies have estimated the overall US excess mortality^2,3^ or excess from individual causes of death at different stages of the pandemic^21–25,28^, our study is the first to examine excess mortality across a full range of causes of death to understand both the direct and indirect mortality costs of the pandemic through the end of 2022.

The pandemic exacerbated already high levels of deaths due to substance use in the US, with relative increases larger than all other non-COVID-19 causes of death across most ages. Social isolation and economic stress may have increased demand for drugs, and access to treatment and harm reduction services was decreased during pandemic restrictions^29^, potentially increasing the lethality of drug use. Disruption in drug supply can also lead to buying from less trusted sources or buying larger quantities at one time, increasing the risk of overdose^30^. A similar increase in drug-related deaths during the pandemic was *not* seen in the UK, including Scotland which has also suffered from a high level of drug mortality in recent years^31^.

In contrast to the increase in drug-related mortality, deaths by suicide generally decreased during the pandemic, despite significant concerns about the impact of social restrictions and economic stress on mental health^23^. This seeming paradox may be related to increased social cohesion during disasters, a phenomenon that has also been seen during wars or after other disasters such as terrorist attacks^32^. Teen suicide also typically decreases during the summer when school is not in session, and this effect was seen during school closures and re-openings during the pandemic^33^. Overall, suicide returned to roughly expected levels by the autumn of 2021. While the absence of measurable surge in suicide deaths was a welcome pandemic surprise, it does not minimize the increases in anxiety and depression that occurred especially for young people during the pandemic^34^. In our age and sex-disaggregated results (Figure 3), decreases in suicide deaths were most noticeable in ages 45-74, with younger and older age groups not seeing much change from expected levels. Overall, the better-than-expected suicide picture during the pandemic highlights the complex social nature of suicidality and provides opportunities for a better understanding of the contexts that can support people at risk.

Deaths due to cardiovascular disease accounted for the largest number of non-COVID-19 excess deaths (115 765 excess deaths) during the pandemic. While not definitive, the strong correspondence of excess CVD mortality with the peaks and troughs of COVID-19 waves (Figure 1) supports the idea that some of these deaths may be underreported COVID-19 deaths. COVID-19 deaths could be misattributed to other causes for a variety of reasons. Testing was less available early in the pandemic and infections may have been undetected. The proportion of deaths at home also increased during the pandemic^35^, reducing the opportunity for a COVID-19 diagnosis. The political polarization of COVID-19 in the US may have contributed to underreporting that varied by geography^36^. In addition to misattributed COVID-19 mortality, increases in CVD mortality may have occurred due to disruptions in routine or emergency health services during COVID-19 peaks^37^ or healthcare avoidance due to fear of infection. COVID-19 infection can also directly damage cardiovascular health and elevate the risk of death for many months after initial infection^38,39^. Increases in substance abuse may also increase risk of CVD death^40^, and the relative increase in CVD deaths was highest for 30–44-year-olds who also saw the biggest relative increases in substance-related deaths (Figure 3). While it is difficult to fully attribute excess CVD deaths to each of these potential causes, more detailed work that takes advantage of differences in COVID-19 infections, healthcare strain, and longer-term CVD impacts across time and geography may help elucidate the relative importance of each mechanism and the implications for the future of cardiovascular disease mortality in the US.

Despite concerns about the impact of health care disruptions on cancer, overall, there was little evidence of excess cancer mortality during the pandemic. Some excess cancer mortality emerged in 2022 for females over age 75 (eTable 4), but not for males. As some cancers are faster growing than others, it will be important to monitor the potential longer-term impacts of delayed cancer screenings or treatments. The pattern of excess cancer deaths at older ages suggests that there may have been some mortality displacement, whereby people close to death from cancer were more likely to die during major COVID-19 waves and therefore did not show up as cancer deaths in the subsequent months.

Our analyses also highlight the substantial burden of pandemic mortality among younger and working-age adults in the US, a pattern that is unusual among high-income countries, where most mortality was concentrated above age 75^14,41^. US adults aged 30-59 suffered over 295,000 excess deaths (Table 1) through the end of 2022, both reflecting and amplifying pre-existing health vulnerabilities in that age group^42^. This burden is especially noticeable in relative increases, for example an 82% increase in 45-59 year-old males due to residual/COVID-19 mortality compared to 34.5% for males over age 75 (eTable 3). The reasons for the younger age distribution of deaths in the US compared to peer countries merits on-going research. While overall excess deaths declined significantly in 2022 compared to 2020 and 2021, they were still substantial (292 629 total deaths, 9.88% above expected; eTable 4), despite the availability of effective COVID-19 vaccines. While COVID-19/residual excess deaths were dramatically lower in the latter half of 2022, excess deaths from other causes including CVD, diabetes and substance use remained significantly elevated (Figure 1). Whether this excess will continue beyond 2022 remains to be seen, but the possibility of a new and higher baseline mortality level is concerning.

### Strengths and Limitations

Our analysis has several limitations. The 2022 US mortality data is still provisional and might be subject to further updates, especially for external causes that require investigation. While we analyzed a comprehensive list of specific causes of death, the computational demands of the GAM model required that we limit to a broad set of categories. Examining other specific causes, such as homicides, Alzheimer disease and related dementias (ADRD), or finer-grained classifications of cardiovascular and metabolic disease, may yield additional insights. Our age-stratified and compositionally robust modeling approach did not allow us to model causes not occurring at all ages, so we were unable to look at ADRD, which Chen, et al^22^ identified as a source of non-COVID-19 excess in the first year of the pandemic. We did not disaggregate excess mortality by geography or race/ethnicity, which have been strongly associated with COVID-19 mortality^16,43^. Further examination of how cause-specific mortality differed by social factors could help illuminate both the direct and indirect pathways through which the pandemic impacted mortality.

A strength of our study is the use of a novel method for calculating compositionally-robust cause-specific excess mortality. Estimating excess single causes independently can lead to unreliable prediction intervals and lack of agreement between cause-specific and total expected deaths^44^. Our model avoids these pitfalls by explicitly accounting for the dependencies between cause-specific deaths and total deaths. We also provide the first full accounting of total and cause-specific US excess mortality by age and sex until the end of 2022, covering the full acute phase of the pandemic. We show both absolute and relative excess mortality estimates, which emphasize different aspects of the pandemic mortality burden, including the disproportionate impact on younger and working age Americans.

The US suffered over 1.3 million excess deaths from March 2020 through the end of 2022. While the majority (73.4%) were attributed to official COVID deaths, the US also saw significant increases in mortality due to substance abuse, cardiovascular disease, diabetes, and accidents. Despite the fall in COVID-specific deaths, the pandemic could have an on-going impact on mortality from other causes that require attention from the medical and public health communities.

## MATERIALS AND METHODS

Monthly death data in the US from 2015 to 2022 came from the Multiple Cause of Death Data via Centers for Disease Control and Prevention’s (CDC) Wonder ^45^. Deaths were stratified by sex and five age groups: 15-29, 30-44, 45-59, 60-74, and 75+. We analyzed eight mutually exclusive causes of death: accidents, cardiovascular diseases (CVD), cancer, diabetes, influenza and pneumonia, substance-related (drugs and alcohol), suicide, and a residual category that included deaths coded as COVID-19. COVID-19 is included in the residual category because it was not an official cause prior 2020 and thus can’t be estimated as separate category for the baseline period. ICD-10 codes corresponding to each category can be found in eTable 1.

We estimated cause-specific excess mortality in the US from March 2020 through December 2022. Using generalized additive^46^ and compositional regression^47^, we separately estimate the expected total death counts and the expected proportions of deaths by cause over year and month which we combine to calculate the cause-specific expected deaths. Such compositional estimation approaches ensure coherency between total and cause-specific expected deaths and explicitly address the dependency among different causes of death^48^. Intuitively, excess mortality counts deaths that occur in the follow-up period above and beyond what would have been expected based on previous trends. For total excess mortality this means that the estimates do not rely on accurate coding of deaths during a crisis (for COVID-19 for example). For cause-specific deaths, this method is more accurate than looking at simple changes over time, because it accounts for pre-existing trends and changes in population age composition over time.

Generalized additive models have proven popular in the excess death modeling literature because of consistently low biases^49^. For the total death counts we employ a conventional specification with monthly fixed-effects and a log-linear long-term trend using a negative-binominal likelihood (see supplementary material).

The cause specific proportions of all deaths also feature seasonality (e.g., more deaths in winter with a higher share of influenza and pneumonia deaths) and long-term trends (e.g., increasing total death counts and an increasing share of substance-related deaths), which we model via monthly coefficients and a linear long term trend in a compositional regression setting using the isometric log-ratio transformation^50^ on the cause of death proportions.

We used data from January 2015 to February 2020 as our baseline data. We predicted the expected number of deaths for each month from March 2020 to December 2022. Excess deaths were calculated as the difference between the observed and expected deaths for each month. Confidence intervals for each cause and age-sex stratification were calculated by iteratively simulating the model 1000 times.

In addition to reporting the absolute difference between expected and observed deaths, we also calculate relative differences, or P-scores. P-scores represent the standardized increase/decrease in percentage difference and are calculated as follows (1):

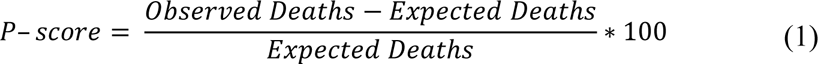

Cumulative P-scores aggregate the monthly standardized differences over a specified time period, offering a consolidated view of the relative difference between observed and expected deaths across an extended duration rather than monthly variations.

All analyses were conducted in R version 4.3.0. Materials for reproduction, including metadata and scripts, will be available at peer review and upon publication.

## Data Availability

Materials for reproduction, including metadata and scripts, will be available at peer review and upon publication.

## Acknowledgments

We acknowledge funding from the Leverhulme Trust (Grant RC-2018-003) for the Leverhulme Centre for Demographic Science (ED, AMT, JD), the European Research Council grant ERC-2021-CoG-101002587 (ED, AMT, JD), the UK Research and Innovation (UKRI) under the UK government’s Horizon Europe funding guarantee EP/X027678/1 (AMT). Previous versions of this manuscript benefited from feedback provided by the Leverhulme Centre for Demographic Science’s Health Inequality Working Group. The content of this manuscript is solely the responsibility of the authors and does not necessarily represent the official views of the ERC, the UKRI, or the Leverhulme Trust.

## Online-Only Supplement

### Technical Material

We used a Generalized Additive Model (GAM). The GAM was chosen due to its versatility in expanding upon the capabilities of Generalized Linear Models (GLMs), accommodating non-linear associations between predictor variables and the response. Specifically, the GAM captures non-linear correlations and fluctuations in extensive datasets through smooth functions, an essential characteristic for addressing seasonality and ensuring precise forecasts.

We used a Poisson distribution. The log link function ensured our model’s predictions were non-negative. This combination of Poisson distribution within the GAM provides a robust tool for understanding temporal variations in death rates.

Our model was defined as (E1):

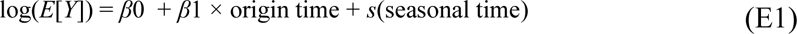

Where *E*[*Y*] is the expected number of deaths and *s* is a smooth function of seasonal time using cubic splines.

### Online-Only Figures and Tables

**eTable 1:** ICD-10 codes corresponding to each cause of death.

| Category | ICD-10 Codes |
| --- | --- |
| Accidents | V00-V99, W00-W99, X00-X59, Y85-Y86 |
| Cancers | C00-D49 |
| CVD | I00-I78 |
| Diabetes | E10-14 |
| Influenza and Pneumonia | J09-J18 |
| Residual | All-cause (minus) [Sum of cause-specific]; includes COVID-19 (U07.1) |
| Substance | F10-F16, F19, K70, K73-K74, X40-45, X85, Y10-Y15 |
| Suicide | U03, X60-X84, Y87 |

**eTable 2.** Cumulative Excess by Cause US, Jan 2020 to Dec 2022.

|  | Median | 95% CI |
| --- | --- | --- |
| Residual | 1 042 445 | (1 018 379; 1 066 754) |
| Accidents | 25 028 | (21 081; 28 832) |
| Cancer | 3 537 | (-4 528; 11 539) |
| CVD | 115 765 | (98 697; 133 783) |
| Diabetes | 41 326 | (38 699; 43 896) |
| Influenza | -8 556 | (-15 236; -2 572) |
| Substance | 86 637 | (79 273; 93 690) |
| Suicide | -8 556 | (-15 236; -2 572) |
| Total | 1 298 763 | (1 226 542; 1 370 804) |

**eTable 3:**
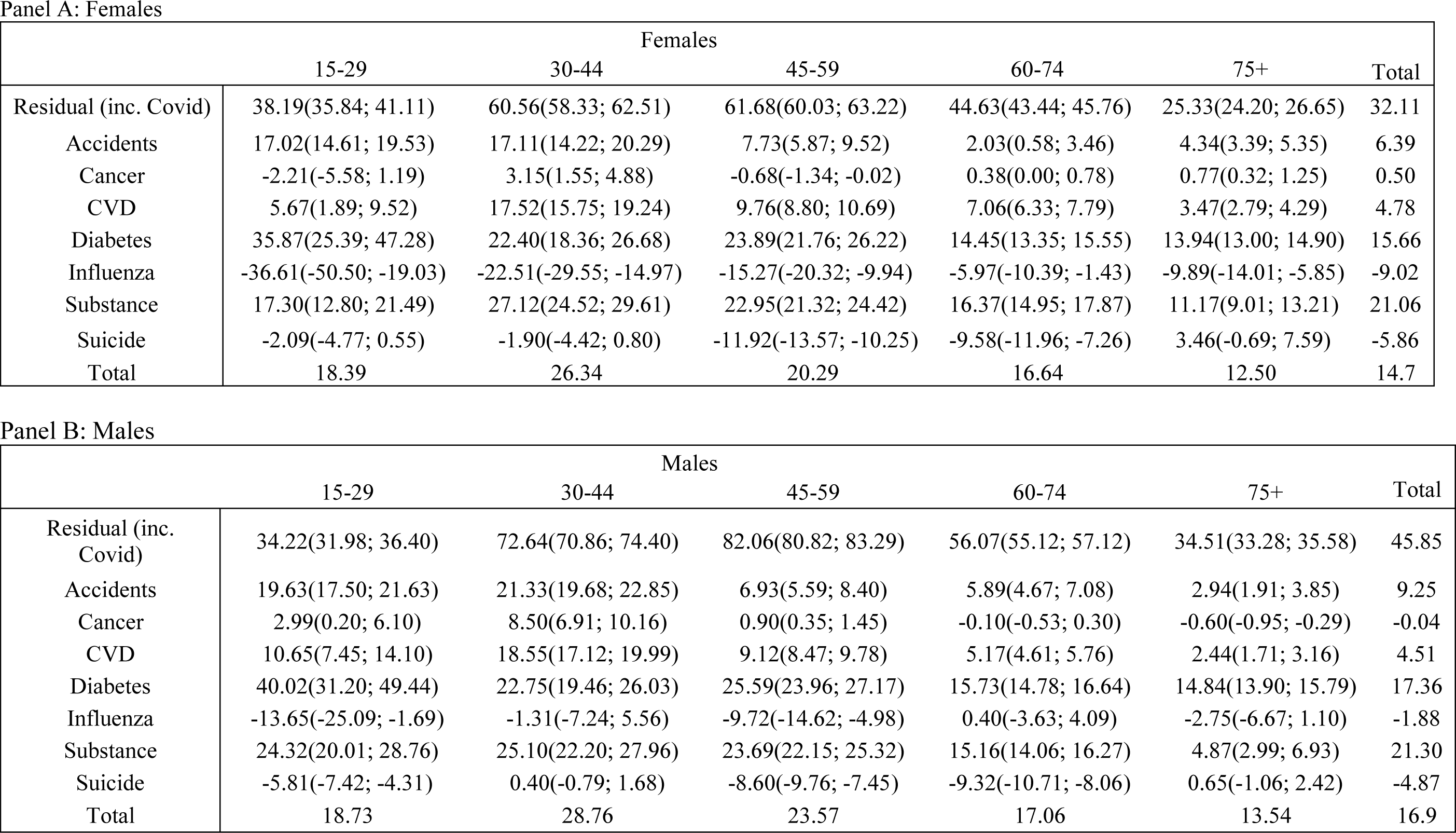
Cumulative Pscores by strata and cause, US, Jan 2020 to Dec 2022for females (Panel A) and males (Panel B)

|  | Females |  |  |  |  |  |
| --- | --- | --- | --- | --- | --- | --- |
|  | 15-29 | 30-44 | 45-59 | 60-74 | 75+ | Total |
| Residual (inc. Covid) | 38.19(35.84; 41.11) | 60.56(58.33; 62.51) | 61.68(60.03; 63.22) | 44.63(43.44; 45.76) | 25.33(24.20; 26.65) | 32.11 |
| Accidents | 17.02(14.61; 19.53) | 17.11(14.22; 20.29) | 7.73(5.87; 9.52) | 2.03(0.58; 3.46) | 4.34(3.39; 5.35) | 6.39 |
| Cancer | -2.21(-5.58; 1.19) | 3.15(1.55; 4.88) | -0.68(-1.34; -0.02) | 0.38(0.00; 0.78) | 0.77(0.32; 1.25) | 0.50 |
| CVD | 5.67(1.89; 9.52) | 17.52(15.75; 19.24) | 9.76(8.80; 10.69) | 7.06(6.33; 7.79) | 3.47(2.79; 4.29) | 4.78 |
| Diabetes | 35.87(25.39; 47.28) | 22.40(18.36; 26.68) | 23.89(21.76; 26.22) | 14.45(13.35; 15.55) | 13.94(13.00; 14.90) | 15.66 |
| Influenza | -36.61(-50.50; -19.03) | -22.51(-29.55; -14.97) | -15.27(-20.32; -9.94) | -5.97(-10.39; -1.43) | -9.89(-14.01; -5.85) | -9.02 |
| Substance | 17.30(12.80; 21.49) | 27.12(24.52; 29.61) | 22.95(21.32; 24.42) | 16.37(14.95; 17.87) | 11.17(9.01; 13.21) | 21.06 |
| Suicide | -2.09(-4.77; 0.55) | -1.90(-4.42; 0.80) | -11.92(-13.57; -10.25) | -9.58(-11.96; -7.26) | 3.46(-0.69; 7.59) | -5.86 |
| Total | 18.39 | 26.34 | 20.29 | 16.64 | 12.50 | 14.7 |

Panel B: Males
|  | Males |  |  |  |  |  |
| --- | --- | --- | --- | --- | --- | --- |
|  | 15-29 | 30-44 | 45-59 | 60-74 | 75+ | Total |
| Residual (inc. Covid) | 34.22(31.98; 36.40) | 72.64(70.86; 74.40) | 82.06(80.82; 83.29) | 56.07(55.12; 57.12) | 34.51(33.28; 35.58) | 45.85 |
| Accidents | 19.63(17.50; 21.63) | 21.33(19.68; 22.85) | 6.93(5.59; 8.40) | 5.89(4.67; 7.08) | 2.94(1.91; 3.85) | 9.25 |
| Cancer | 2.99(0.20; 6.10) | 8.50(6.91; 10.16) | 0.90(0.35; 1.45) | -0.10(-0.53; 0.30) | -0.60(-0.95; -0.29) | -0.04 |
| CVD | 10.65(7.45; 14.10) | 18.55(17.12; 19.99) | 9.12(8.47; 9.78) | 5.17(4.61; 5.76) | 2.44(1.71; 3.16) | 4.51 |
| Diabetes | 40.02(31.20; 49.44) | 22.75(19.46; 26.03) | 25.59(23.96; 27.17) | 15.73(14.78; 16.64) | 14.84(13.90; 15.79) | 17.36 |
| Influenza | -13.65(-25.09; -1.69) | -1.31(-7.24; 5.56) | -9.72(-14.62; -4.98) | 0.40(-3.63; 4.09) | -2.75(-6.67; 1.10) | -1.88 |
| Substance | 24.32(20.01; 28.76) | 25.10(22.20; 27.96) | 23.69(22.15; 25.32) | 15.16(14.06; 16.27) | 4.87(2.99; 6.93) | 21.30 |
| Suicide | -5.81(-7.42; -4.31) | 0.40(-0.79; 1.68) | -8.60(-9.76; -7.45) | -9.32(-10.71; -8.06) | 0.65(-1.06; 2.42) | -4.87 |
| Total | 18.73 | 28.76 | 23.57 | 17.06 | 13.54 | 16.9 |

Panel A: females

**eTable 4:** Cumulative Excess by strata, cause, and year, US Jan 2020 to Dec 2022 for females (Panel A) and males (Panel B) Panel A: females.

|  | 15-29 |  |  | 30-44 |  |  | 45-59 |  |  | 60-74 |  |  | 75+ |  |  | Total |  |  |
| --- | --- | --- | --- | --- | --- | --- | --- | --- | --- | --- | --- | --- | --- | --- | --- | --- | --- | --- |
|  | 2020 | 2021 | 2022 | 2020 | 2021 | 2022 | 2020 | 2021 | 2022 | 2020 | 2021 | 2022 | 2020 | 2021 | 2022 | 2020 | 2021 | 2022 |
| Residual (incl. Covid) | 1,134 | 2,269 | 1,179 | 3,700 | 9,816 | 3,500 | 13,547 | 28,064 | 9,616 | 42,004 | 63,801 | 26,367 | 127,490 | 81,476 | 58,802 | 187,875 | 185,426 | 99,464 |
| Accidents | 383 | 689 | 482 | 337 | 751 | 560 | 169 | 421 | 331 | -62 | 346 | 181 | 217 | 1,101 | 1,549 | 1,044 | 3,308 | 3,103 |
| Cancer | -5 | -57 | 1 | -72 | 382 | 386 | -300 | -303 | -95 | 292 | 1,180 | -240 | -448 | 226 | 3,193 | -533 | 1,428 | 3,245 |
| CVD | 85 | 79 | -2 | 706 | 1,069 | 923 | 2,248 | 2,853 | 1,784 | 5,131 | 6,977 | 4,671 | 12,239 | 4,514 | 13,441 | 20,409 | 15,492 | 20,817 |
| Diabetes | 53 | 106 | 83 | 237 | 295 | 180 | 882 | 1,161 | 894 | 2,073 | 2,117 | 1,203 | 3,081 | 2,383 | 2,293 | 6,326 | 6,062 | 4,653 |
| Influenza | -5 | -46 | -50 | -13 | -205 | -131 | 73 | -468 | -352 | 468 | -935 | -347 | 40 | -3,566 | -559 | 563 | -5,220 | -1,439 |
| Substance | 787 | 947 | 444 | 2,439 | 3,900 | 2,718 | 2,267 | 4,030 | 3,022 | 1,093 | 2,329 | 1,726 | 271 | 393 | 566 | 6,857 | 11,599 | 8,476 |
| Suicide | -64 | 16 | -55 | -157 | -86 | 125 | -434 | -354 | -235 | -221 | -202 | -105 | -33 | 9 | 90 | -909 | -617 | -180 |
| Total | 2,368 | 4,003 | 2,082 | 7,177 | 15,922 | 8,261 | 18,452 | 35,404 | 14,965 | 50,778 | 75,613 | 33,456 | 142,857 | 86,536 | 79,375 | 221,632 | 217,478 | 138,139 |

Panel B: males
|  | 15-29 |  |  | 30-44 |  |  | 45-59 |  |  | 60-74 |  |  | 75+ |  |  | Total |  |  |
| --- | --- | --- | --- | --- | --- | --- | --- | --- | --- | --- | --- | --- | --- | --- | --- | --- | --- | --- |
|  | 2020 | 2021 | 2022 | 2020 | 2021 | 2022 | 2020 | 2021 | 2022 | 2020 | 2021 | 2022 | 2020 | 2021 | 2022 | 2020 | 2021 | 2022 |
| Residual (incl. Covid) | 3,295 | 5,024 | 2,604 | 8,346 | 17,388 | 6,882 | 25,848 | 47,224 | 14,429 | 67,397 | 89,687 | 33,258 | 108,660 | 86,658 | 53,886 | 213,546 | 245,981 | 111,059 |
| Accidents | 1,277 | 1,998 | 1,714 | 1,214 | 2,561 | 2,217 | 441 | 1,173 | 754 | 270 | 1,294 | 1,067 | 30 | 660 | 1,155 | 3,232 | 7,686 | 6,907 |
| Cancer | -47 | 62 | 125 | 222 | 490 | 645 | 71 | 209 | 686 | 130 | 954 | -1,469 | -704 | -1,414 | -364 | -328 | 301 | -377 |
| CVD | 115 | 281 | 115 | 1,570 | 2,354 | 1,766 | 5,354 | 5,901 | 2,771 | 8,799 | 9,894 | 3,204 | 9,760 | 1,510 | 6,201 | 25,598 | 19,940 | 14,057 |
| Diabetes | 108 | 170 | 107 | 414 | 453 | 360 | 1,756 | 2,072 | 1,737 | 3,168 | 3,682 | 2,012 | 2,887 | 2,639 | 2,763 | 8,333 | 9,016 | 6,979 |
| Influenza | 3 | -25 | -20 | 86 | -61 | -15 | 272 | -395 | -505 | 1,084 | -515 | -392 | 1,040 | -1,700 | -171 | 2,485 | -2,696 | -1,103 |
| Substance | 2,780 | 3,016 | 1,538 | 5,510 | 8,563 | 6,148 | 5,428 | 9,233 | 6,804 | 2,369 | 4,666 | 3,572 | 145 | 183 | 413 | 16,232 | 25,661 | 18,475 |
| Suicide | -343 | -23 | -1,078 | -247 | 172 | 214 | -1,070 | -931 | -323 | -822 | -668 | -501 | 11 | -104 | 181 | -2,471 | -1,554 | -1,507 |
| Total | 7,188 | 10,503 | 5,105 | 17,115 | 31,920 | 18,217 | 38,100 | 64,486 | 26,353 | 82,395 | 108,994 | 40,751 | 121,829 | 88,432 | 64,064 | 266,627 | 304,335 | 154,490 |

**eFigure 1:**
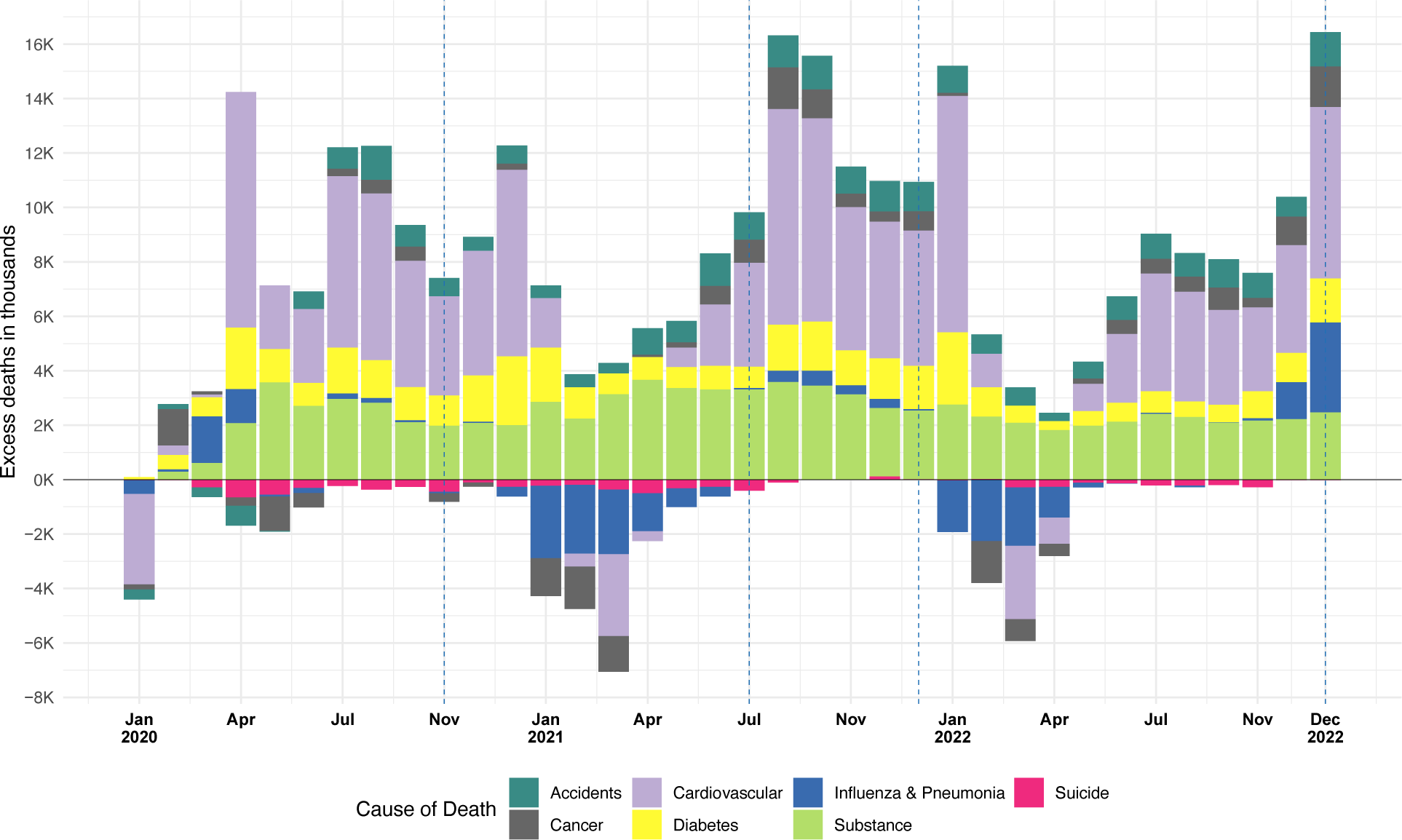
Absolute excess mortality by cause of death (without residual/COVID), US, Jan 2020 to Dec 2022.

**eFigure 2:**
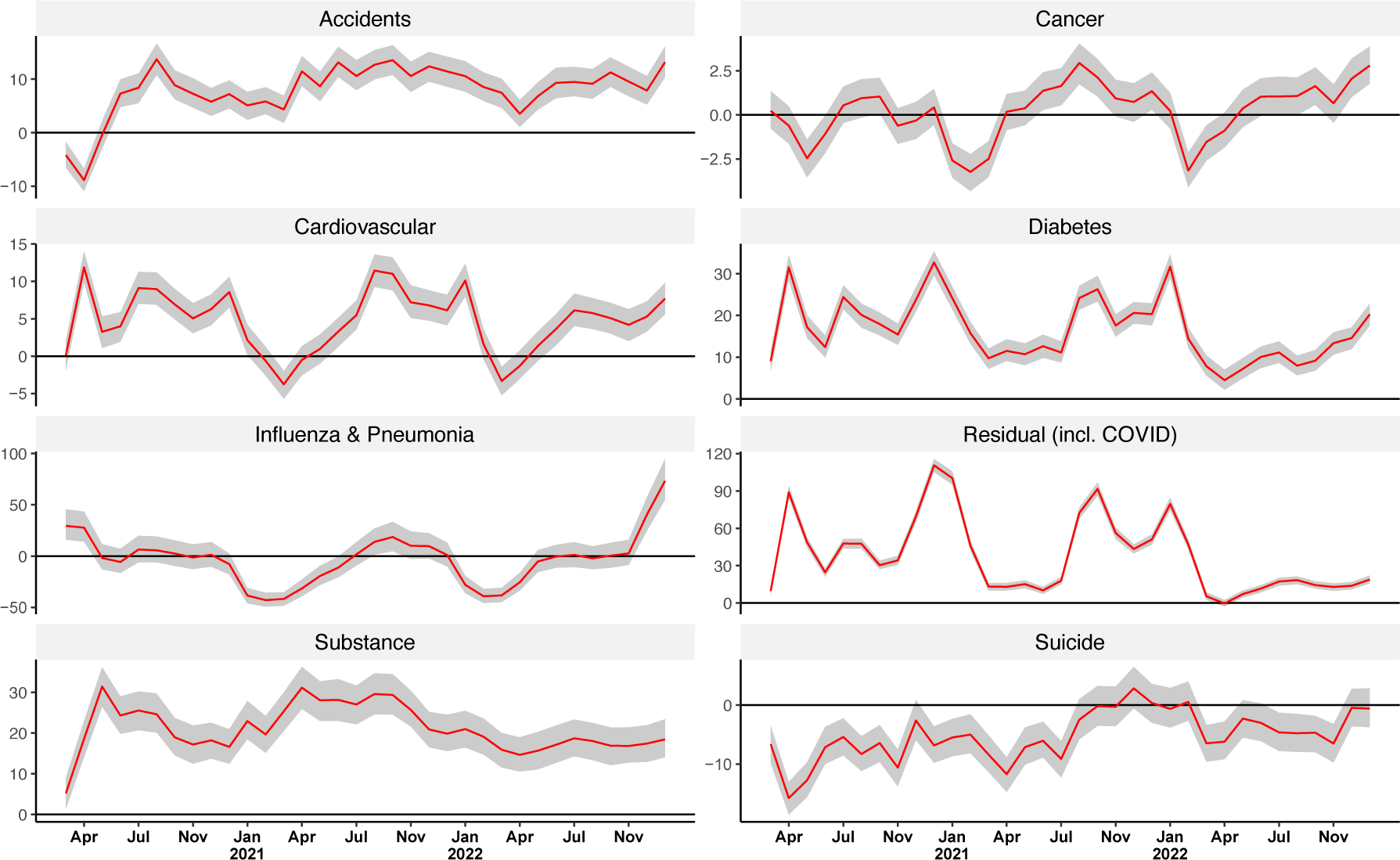
P-scores by cause of death, ages 15+, US, Mar 2020 to Dec 2022. Notes. Data come from the Centers for Disease Control and Prevention (CDC) Wonder and authors’ own estimates.

